# Method for Active Pandemic Curve Management (MAPCM)

**DOI:** 10.1101/2020.04.06.20055699

**Authors:** Willem G. Odendaal

## Abstract

The COVID-19 pandemic of 2020 prompted stringent mitigation measures to “flatten the curve” quickly leading to an asphyxiated US economy as a national side effect. There are severe drawbacks to this strategy. The resulting *flattened curve* remains exponential and always under utilizes available healthcare capacity with a chance of still overburdening it. Moreover, while a mitigation strategy involving isolation and containment can scale down infections, it not only prolongs the outbreak significantly, but also leaves a susceptible population in its wake that’s ripe for a secondary outbreak. Since economic activity is inversely proportional to mitigation, curtailing the outbreak with sustained mitigation can stifle the economy severely with disastrous repercussions. Full mitigation for the duration of an outbreak is therefore unsustainable and, overall, a poor solution with potentially catastrophic consequences that could’ve been avoided. A new strategy, coined a *“Method for Active Pandemic Curve Management”*, or *MAPCM*, presented herein can shape the outbreak curve in a controlled manner for optimal utilization of healthcare resources during the pandemic, while drastically shortening the outbreak duration compared to mitigation by itself without trading off lives. This method allows mitigation measures to be relaxed gradually from day one, which enables economic activity to resume gradually from the onset of a pandemic. Since outbreak curves (such as hospitalizations) can be programmed using this method, they can also be shaped to accommodate changing needs during the outbreak; and to build herd immunity without the damaging side effects. The method can also be used to ease out of containment. **MAPCM is a method and not a model**. It is compatible with any appropriate outbreak model; and herein it is illustrated in examples using a hybrid logistic model.

## I. Introduction

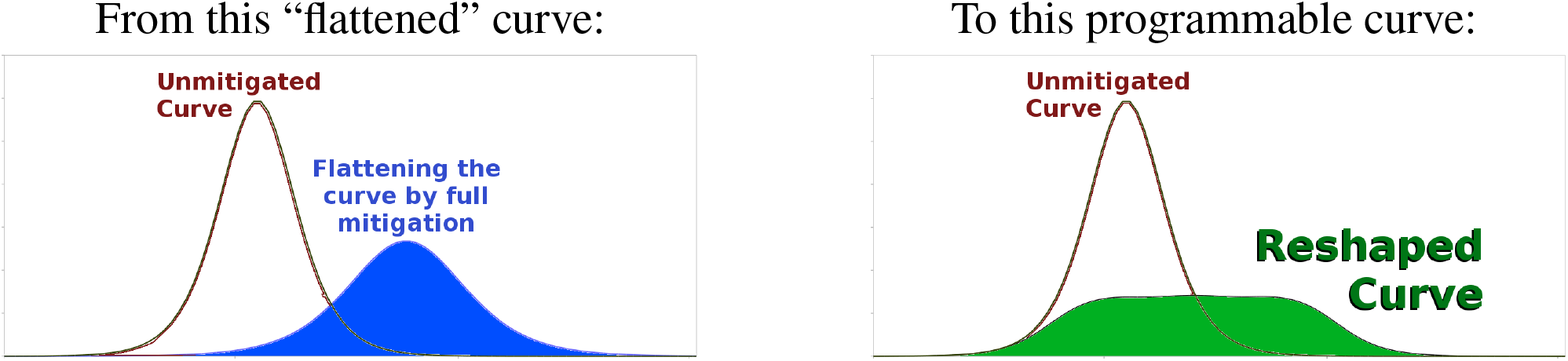

### Being behind the curve

#### Strengths

- slows down the outbreak
- reduces infection rate (the total percentage of population infected)
- shrinks (scales down) the original curve
- buys time (seasonal damping, finding cure, preparing healthcare, etc.)
- potentially reduces fatality rate

#### Remaining Weaknesses

- remains exponential
- still has a high peak
- can still overburden available healthcare capacity around its peak
- under utilizes available healthcare capacity for long periods before and after peak
- still costs too many lives that could have been saved
- prolongs the outbreak for too long
- high cost of containment for entire country
- unsustainable - economy frozen and in free fall

### Being controlled by the outbreak

### Getting ahead of the curve

#### Pros

- programmable curve in amplitude and duration
- can be customized to meet available resources
- eliminates the exponential peak
- optimizes utilization of healthcare capacity
- reduces fatality rate through better healthcare
- shortens outbreak duration & disruption to normal life
- minimizes the cost of containment
- does not keep economy at standstill
- gradually brings economy back online from day one
- can build herd immunity safely
- saves lives

#### Cons

- has never been done for a pandemic before
- uncertainties due to stochastics and the human factor

### Controlling the outbreak

When the COVID-19 pandemic broke out, it soon became clear that the healthcare systems in possibly every country in the world would be crippled by an overwhelming need for hospitalization exceeding the available resources and supply chains. While many vital characteristics of the pathogen are still unclear due to lacking data, it appears that the virus may not be as deadly as it is contagious. The actual fatality rate and infection rates haven’t fully crystallized yet due to insufficient testing and a lack of population studies that should’ve been conducted months earlier. Even if the fatality rate is close to that of influenza, this virus distinguishes itself by concentrating annual totals for deaths and critical care needs into a matter of weeks if there is no intervention. A worrying statistic is the death rate among resolved cases, which varies between 10% to over 90%. However, these figures appear to be dwarfed by all the infections that remained largely asymptomatic or undetected.

The most common strategy implemented by governments to address these problems came down to aggressive containment, sometimes borderlining draconian laws to *“flatten the curve”*. The idea is to slow down the spread, reduce the infection rate, and close the gap between the excessive demand for healthcare and the available resources. However, the level of mitigation required to flatten the curve can suffocate an economy due to layoffs and closing businesses. To make matters worse, sustained mitigation also prolongs the outbreak for an unsustainable amount of time and leaves the population suceptible to a secondary outbreak. Moreover, the *“flattened curve”* always under utilizes available healthcare resources for long periods of time and might still exceed the threshold during the peak, as the curve maintains its exponential character despite being damped.

The question then becomes whether it is possible to shrink the gap between hospitalizations and available healthcare capacity while keeping healthy economic activity without sacrificing lives. In this paper it will be shown that not only is this possible, but the duration of the outbreak can also be shortened, loss of life minimized, and healthcare utilization optimized. All this is possible using a new Method for Active Pandemic Curve Management (MAPCM). It is accomplished by considering the problem from a systems engineering perspective and implementing a quasi-open-loop control scheme using existing mitigation mechanisms to curb the pandemic outbreak in a controlled way. MAPCM makes it possible to surgically program the curve to the point of both meeting changing needs and safely building herd immunity in a controlled fashion.

MAPCM will be counter-intuitive for most people, especially those in the healthcare professions. However, equally counter-intuitive control methods do exist in existing electrical, industrial, and military products and applications.

Please note that **the focus of this paper is to present a new method, not a model**. The models and numbers used in the rest of the paper are unimportant and can be substituted by any other suitable models, simulations, or other constructs. For illustration purposes, a hybrid logistic model for the COVID-19 pandemic will be applied to the incoming case statistics in USA by way of a few examples.

Also note that **the reproductive number**, *R*_*o*_, is defined in this paper as the number of people infected per person per day:

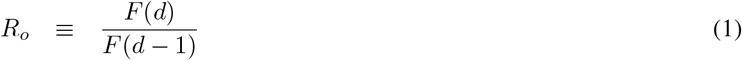

where *F* (*d*) is the relevant cumulative figure on day *d* during the initial stages of the outbreak.

## II. Shortcomings of Aggressive Containment Strategies

### A. Economic Impact

Travel bans and aggressive containment strategies did have an effect on slowing down the spreading of COVID-19. Since the outbreak caught the entire world except China by surprise, the ability to buy some time by imposing mitigation measures is certainly desirable, especially in view of the absence of any useful information about the disease. However, the cost of aggressive mitigation for even a short amount of time is stupendous. The stock market plunged within a day of announcing even mild mitigation through voluntary self-isolation in the United States. Closing business for all but essential functions lead to layoffs and skyrocketing unemployment that will have serious consequences for any country. It quickly became apparent that these outbreak mitigation strategies:

1. are inversely proportional to economic health (see Figure 1),
2. are extremely costly, and
3. suspending economic activity over even brief periods of time can sharply spike unemployment, business closures, widespread panic, suicides, etc.

Sustained mitigation to flatten the curve is the worst of both worlds from an economic perspective as shown in Figure 1(a). Not only is the economic activity halted, thus pushing an otherwise healthy economy into freefall, it also prolongs it well beyond the unmitigated rate of gaining herd immunity situation. As shown in Figure 1(b) MAPCM begins with similar levels of mitigation, but immediately starts lifting them gradually, which eases the economy back to normal from day one.

**Fig. 1.**
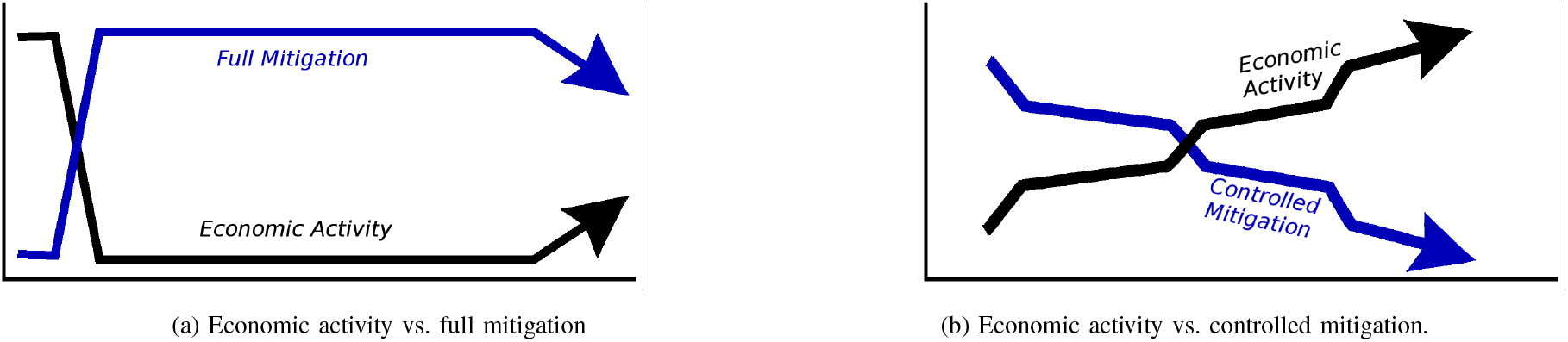
Economic Health and mitigation measures are inversely proportional

### B. Healthcare Utilization

A disappointing outcome of sustained full mitigation by means of stringent containment strategies is that, while the outbreak curves are indeed being *scaled down*, they aren’t *flattened* by much. In fact, the curves for projected hospitalizations with full mitigation remain exponential with peak amplitudes that may still exceed available healthcare capacities as shown in Figure 2 and again in Figure 3, which depicts new cases being reported daily under a few different strategies.

**Fig. 2.**
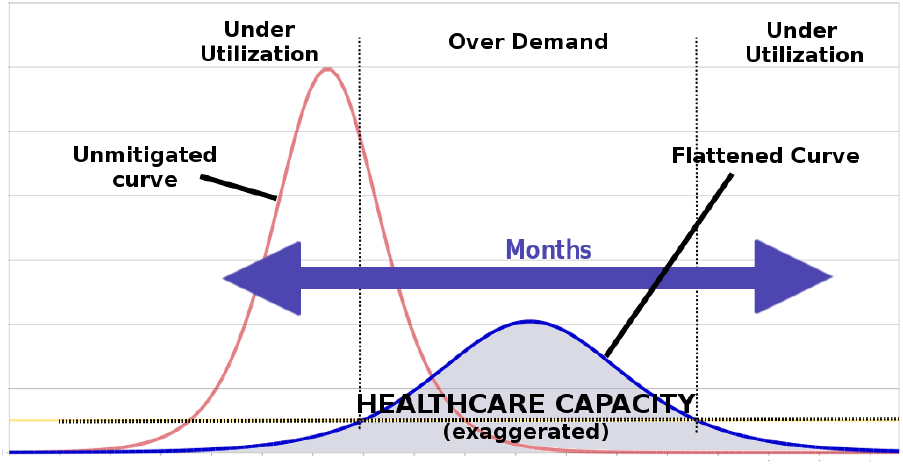
Shortcomings of the “flattened” curve by sustained mitigation

**Fig. 3.**
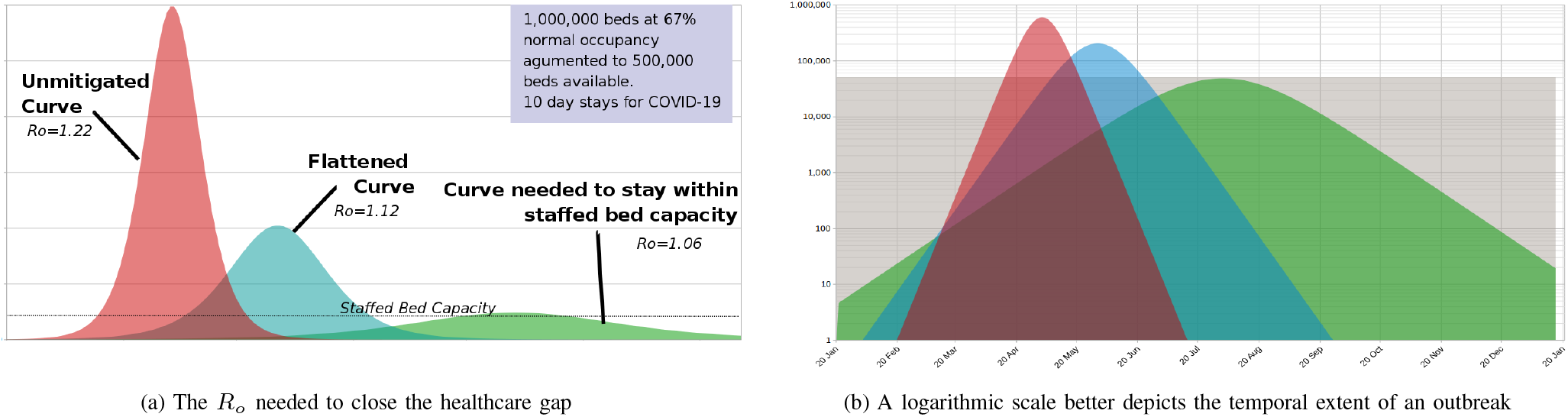
Mitigation level vs. viral spread as function of *R*_*o*_

In Figure 2 the *“flattened”* curve using sustained mitigation still overwhelms available staffed beds during and around the peak period. However, notice that the curve has two long tail ends in the front and rear during which the available healthcare resources are being severely underutilized. In this example a normalized *reproductive number*^1^, *R*_*o*_ = 1.22, was used (based on a curve-fit from the USA data before a national emergency was declared.)

Let’s call the curve with *R*_*o*_ = 1.22 the unmitigated curve for USA, with an assumed infection rate of approximately 18%. *Infection rate* is simply the percentage of the total population who will be infected. After the emergency was declared and mitigation strategies implemented, a new *R*_*o*_ = 1.12 was obtained (from an updated curve fit) for which we’ll assume an infection rate of 11%. Assuming a hospitalization rate of 20% for COVID-19, Figure 3 shows what the original *unmitigated* (*R*_*o*_ = 1.22) and *flattened* (*R*_*o*_ = 1.12) curves for hospitalizations look like next to one another. It also shows what the curve would look like if *R*_*o*_ = 1.06 to force the peak of the curve match available resources.

### C. Outbreak Duration

Notice that the y-axis in Figure 3(a) goes from zero to very large numbers, so that numbers of tens, hundreds, or thousands are indistinguishably small. Yet a small number of infections can lead to an outbreak or a secondary wave. It is therefore a poor graphical representation for estimating the duration of an outbreak. To estimate how long an outbreak might last given a mitigation strategy, it is better to use a logarithmic scale as depicted in Figure 3(b) which contains exactly the same information as Figure 3(a). It is clear from the figure that no matter which level of sustained mitigation is implemented, the duration of containment is far too long to halt an economy. In each case available healthcare capacity is abysmally underutilized. Only the curve with *R*_*o*_ = 1.06 didn’t exceed the healthcare capacity threshold.

The actual figures for the USA are staggering. There are roughly one million staffed hospital beds [1], [2] in the country, of which about 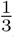 are available on a good day (prior to COVID-19). If the available capacity can be raised to 500,000 for the epidemic, and the average stay of a hospitalized COVID-19 inpatient is 10 days, then on average only 50,000 beds are available per day for new COVID-19 patients.

To remain below this capacity threshold in terms of available staffed beds, the pandemic has to last longer than the *minimum pandemic period, T*_*P*min_. Assuming an infection rate of 18% and a hospitalization rate of 20% then :

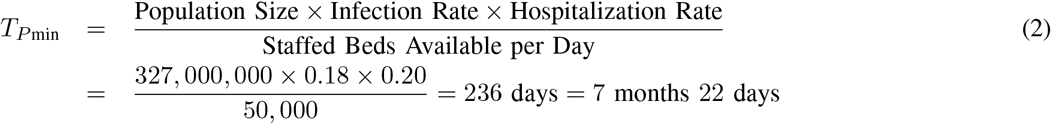

where *infection rate* is simply the percentage of the total population who will be infected and the *hospitalization rate* is the percentage of all infected people who will be hospitalized.

For an infection rate of 11% with the same hospitalization rate of 20%, the idealized *pandemic period* becomes:

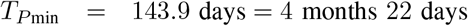

These are the shortest possible durations to remain below the capacity threshold under ideal circumstances in this example. They can be plotted on graphs such as in Figure 3 as an *ideal curve* in the shape of a rectangle. To reduce *T*_*P*min_ without exceeding capacity, either the infection rate, hospitalization rate, or the average stay per patient has to decrease, or the number of staffed beds have to increase.

The important takeaways from this is:

- the national economy cannot endure being shut down even for the period under the ideal curve,
- the period of time under the *flattened curve* is much longer than the length of time under the ideal curve, and
- sustained full mitigation is therefore a very uneffective strategy.

## III. The Concept of Curve Shaping

One of the goals of MAPCM is to eliminate the long *“tails”* of underutilization that form part of the curve. This can be done by reshaping the curve so that the excess of expected inpatients during and around the peak can be treated beforehand or afterwards to fill up the underutilized portions above the curve and under the capacity threshold. This aim of the method is illustrated graphically in Figure 4:

**Fig. 4.**
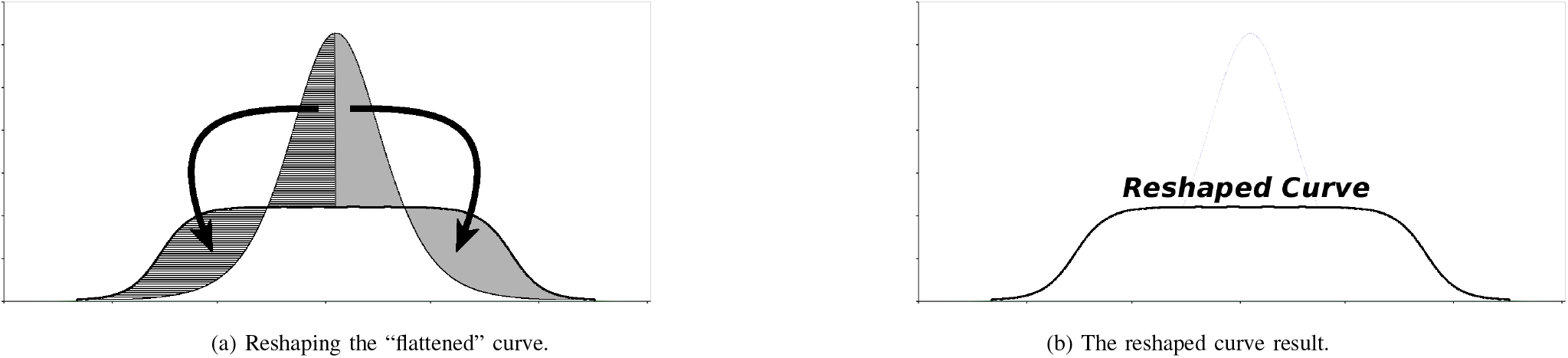
The aim of the method explained graphically

Before explaining how this can be accomplished, and since the pandemic is ongoing, it is necessary to offer a word of warning.

**<u>CAUTION</u>**: The method described in this paper can theoretically produce all the desirable results. However, if the method is either misunderstood or implemented incorrectly, then it is possible to get unexpected results. It is sensitive to timing, amplitudes, segmentation rules, mitigation mechanisms, and other nuances. If you are considering this method, please do not attempt it in practice unless your team has fully explored the method and the underlying mechanisms at play.

– Contact for assistance.

### A. Simplified Example

#### 1) Assumptions

To illustrate the method in its simplest form, let’s assume we have a homogeneous population that can be divided equally in *N*_*s*_ isolated segments that are also homogeneous. Next let’s assume that there is only one possible reproductive number for the virus, *R*_*o*_. For example, it might correspond to the unmitigated rate at which herd immunity would occur. Finally, we’ll assume that there are only two possible states that can be present in each of the segments. In the first state the segment is in complete isolation and the virus has not been introduced into the segment population. In the second state, the virus spreads with *R*_*o*_. If isolation of a segment is terminated, that segment will be infected at *R*_*o*_ up to an *infection rate*^2^, *I*_%_.

#### 2) Initial Conditions

Sufficient containment measures are put in place for the entire population to be in the uninfected, isolation state (or mitigated state).

#### 3) Available Control Mechanism

As the government of the population, we are able to relax isolation measures for any segment(s) on any day of our choosing.

#### 4) Other Rules and Limitations

We are only able to switch each segment from one state to the other once or not at all. A segment cannot be switched back to the original state.

#### 5) Specific Case Numbers

The values for important parameters in this example and the results are given in Figure 5: The curves for the various segments add up to the reshaped curve. The unmitigated curve for herd immunity can be replaced by a controllable, truly flat curve that’s been stretched out over a longer period of time. In fact, by choosing the times and segment sizes differently, the resulting curve’s shape can be programmed to follow a wide variety of custom contours.

**Fig. 5.**
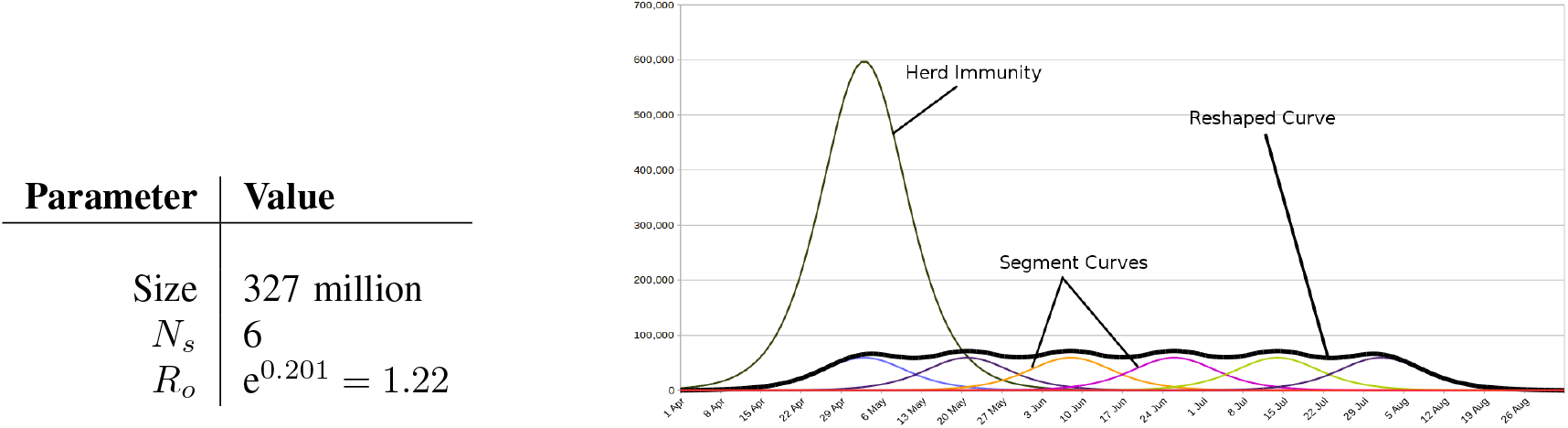
A simplified example to illustrated the concept.

This example serves to illustrates the principle. However, it is idealistic and unrealistic, because segments cannot be isolated perfectly in practice.

## IV. Examples of the Curve Management Method

This section will explore examples that are less idealistic. Keep in mind that these are examples for illustrating the principle based on simple modeling of averages. In practice, pandemic outbreaks are stochastic, irregular, littered with exceptions (i.e. super spreading,) and generally more complex. In all the examples to follow, the method is applied to a logistic function model that was fitted to US data of confirmed cases; and implementation starts between 2 March and 7 March 2020.

### A. Moderate Mitigation

#### 1) Assumptions

In this example let’s assume a homogeneous population that can be divided in *N*_*s*_ isolated segments of varying sizes. Next let’s assume that there are exactly two possible reproductive numbers for the virus, *R*_*oh*_ and *R*_*om*_, where *R*_*oh*_ > *R*_*om*_. For example, *R*_*oh*_ might be the reproductive rate at which unmitigated herd immunity occurs, while *R*_*om*_ is the reproduction number with certain mitigation measures in place. Let’s also assume that there are only two possible states to choose from and that only one of them can be present in each of the segments. In the initial state the virus infects the population of each segment with *R*_*om*_. Once mitigation is relaxed in a specific segment, it switches to the unmitigated herd state having *R*_*oh*_ as the reproductive number.

#### 2) Initial Conditions

Sufficient containment measures are put in place for the entire population to be in the mitigated state, with *R*_*o*_ = *R*_*om*_.

The reproductive number ratio is defined as:

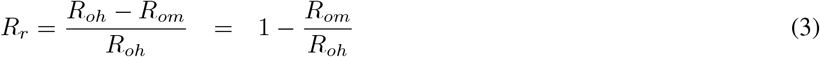

This ratio should increase proportional to the stringency of containment and isolation measures.

#### 3) Available Control Mechanism

As the governing body of the population, we are able to relax mitigation on any segment on any day. Each segment will be infected at reproductive rate *R*_*om*_ until mitigation for that segment is relaxed and it falls back to the higher reproductive number, *R*_*oh*_.

#### 4). Infection Rate

If left at the initial mitigated state, the virus can infect the population of a segment to a maximum of *I*_%*m*_ of the particular segment’s population. If mitigation measures are lifted so *R*_*oh*_ takes effect, the virus can infect the segment’s population to a maximum of *I*_%*h*_. In general, *I*_%*h*_ > *I*_%*m*_. In the example it is assumed that 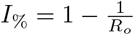 for illustration.

#### 5) Other Rules and Limitations

We are only able to switch each segment once or not at all. Once at *R*_*oh*_, we’ll also assume the rate cannot be switched back to *R*_*om*_. (This is merely to frame the example, and may not be true in practice.)

**TABLE I.** Parameters for example with moderate reproductive ratio.

| Parameter | Value |
| --- | --- |
| Size | 327 million |
| $R_{oh}$ | $e^{0.201} = 1.22$ |
| $R_{om}$ | $e^{0.115} = 1.12$ |
| $R_r$ | 8.2% |
| $I_{\%h}$ | 18% |
| $I_{\%m}$ | 11% |

Figure 6 shows the results^3^ for two examples using moderate mitigation and *R*_*r*_ = 8.2%. With curve management implemented, the mitigation measures in each of the segments are relaxed at precise intervals. The curves for each segments then add up to the thick black curve.

**Fig. 6.**
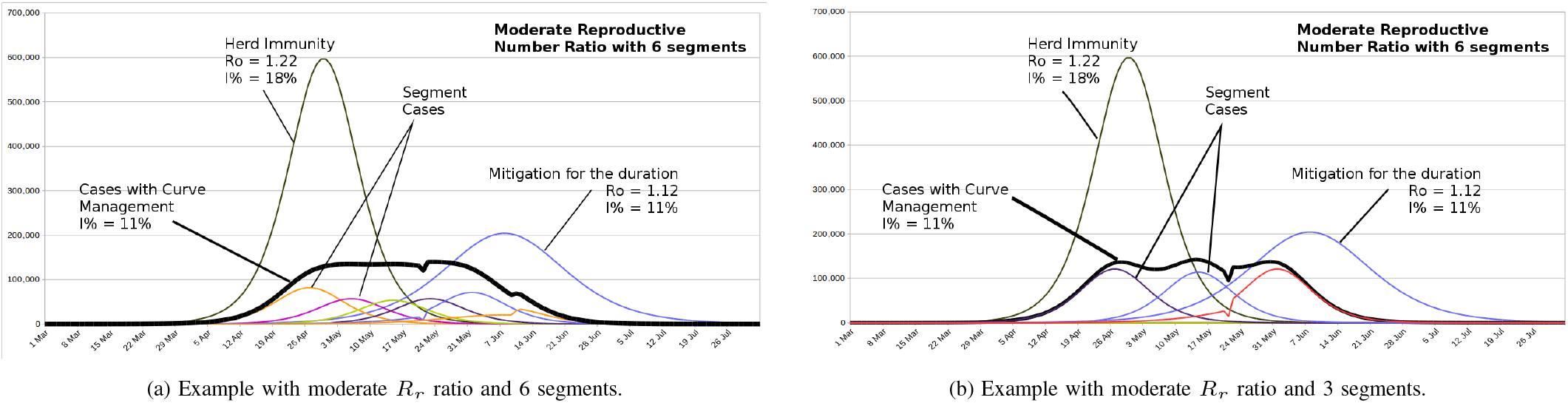
Examples for moderate *R*_*r*_ with 6 and 3 segments showing curves for unmitigted herd-immunity, mitigated-, and controlled infections. Infection rates are limited to *I*_%*m*_.

Figure 7 shows that when *I*_%_ changes from *I*_%*m*_ to *I*_%*h*_ in the segments as mitigations are relaxed, there is no overall improvement over pure mitigation at *R*_*om*_, but still an improvement over the unmitigated curve. The area under the reshaped curve is identical to the area under the unmitigated curve if mitigation in all segments were relaxed at some point.

**Fig. 7.**
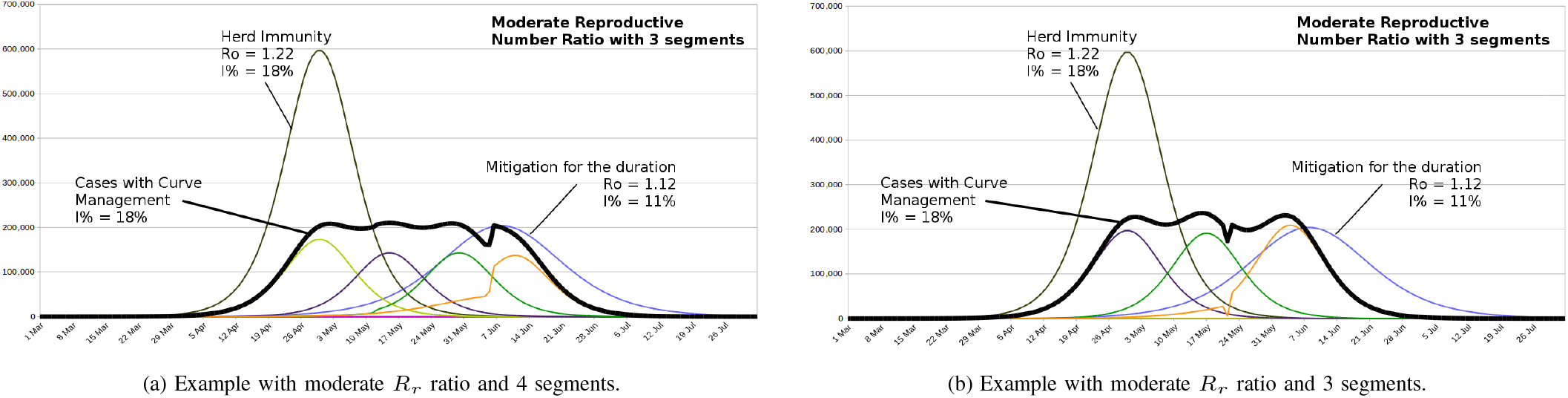
Examples for moderate *R*_*r*_ with 6 and 3 segments showing curves for unmitigated herd-immunity, mitigated-, and controlled infections. Infection rates go up to *I*_%*h*_.

These examples illustrate that, as long as *I*_%_ ≤ *I*_%*h*_:

1. the rate of unmitigated herd immunity is the worst case scenario with the highest peak magnitude,
2. even if the infection rate increases to that of unmitigated herd immunity, the peak of the MAPCM curve can be maintained well below that of sustained mitigation for the duration of the outbreak, and
3. even poor curve management is better than no curve managament at all.

### B. Stringent Mitigation Measures

#### 1) Assumptions

In this example let’s raise *R*_*r*_. Again assume a homogeneous population that can be divided in *N*_*s*_ homogeneous segments of varying sizes. Next let’s assume that there are exactly three possible reproductive numbers for the virus, *R*_*oh*_, *R*_*om*_, and *R*_*os*_ where *R*_*oh*_ > *R*_*om*_ > *R*_*os*_. For example, *R*_*oh*_ might be the rate at which unmitigated herd immunity occurs, *R*_*om*_ the reproduction number with moderate mitigation measures in place, and *R*_*os*_ when stringent mitigation measures are imposed. Finally, we’ll assume that there are only three possible states that each of the segments can be in. In the initial state the virus infects the population of each segment with *R*_*os*_. Once mitigation is relaxed to moderate levels in specific segment, it switches to the unmitigated state with *R*_*o*_ = *R*_*h*_.

#### 2) Initial Conditions

Sufficient containment measures are put in place for the entire population to be in the mitigated state, with *R*_*o*_ = *R*_*os*_.

#### 3) Available Control Mechanism

As the governing body of the population, we are able to relax mitigation on any segment on any day. Each segment will be infected at rate *R*_*os*_ until mitigation for that segment is relaxed and it falls back to the higher infection rate, *R*_*oh*_.

#### 4) Infection Rate

If left at the initial mitigated state, the virus can infect the population of a segment to a maximum of *I*_%*s*_ of the particular segment’s population. If mitigation measures are lifted so *R*_*oh*_ takes effect, the virus will infect the segment’s population to a maximum of *I*_%*h*_ or *I*_%*m*_. In general, *I*_%*h*_ > *I*_%*m*_ > *I*_%*s*_. Again it is assumed that 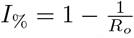 for illustration.

#### 5) Other Rules and Limitations

We are only able to switch each segment once or not at all. Once at *R*_*oh*_, the rate cannot be switched back to *R*_*os*_. (Again, this is merely to frame the example, and is not true in practice.)

**TABLE II.** Parameters for examples with stringent mitigation measures.

| Parameter | Value |
| --- | --- |
| Size | 327 million |
| $R_{oh}$ | $e^{0.29} = 1.34$ |
| $R_{om}$ | $e^{0.19} = 1.21$ |
| $R_{os}$ | $e^{0.10} = 1.11$ |
| $I_{\%h}$ | 25% |
| $I_{\%m}$ | 17.3% |
| $I_{\%s}$ | 9.5% |
| $R_r$ | 20.7% |

Figure 8 shows the results for examples in which the method is applied using these three reproductive numbers. In Figure 8(a) the infection rate can go up to *I*_%*m*_ and in Figure 8(b) to *I*_%*h*_. The need for the more stringent mitigation measures were to buy more time to reduce the amplitude of the controlled curve, which cannot go beyond the curve with *R*_*os*_ time wise.

**Fig. 8.**
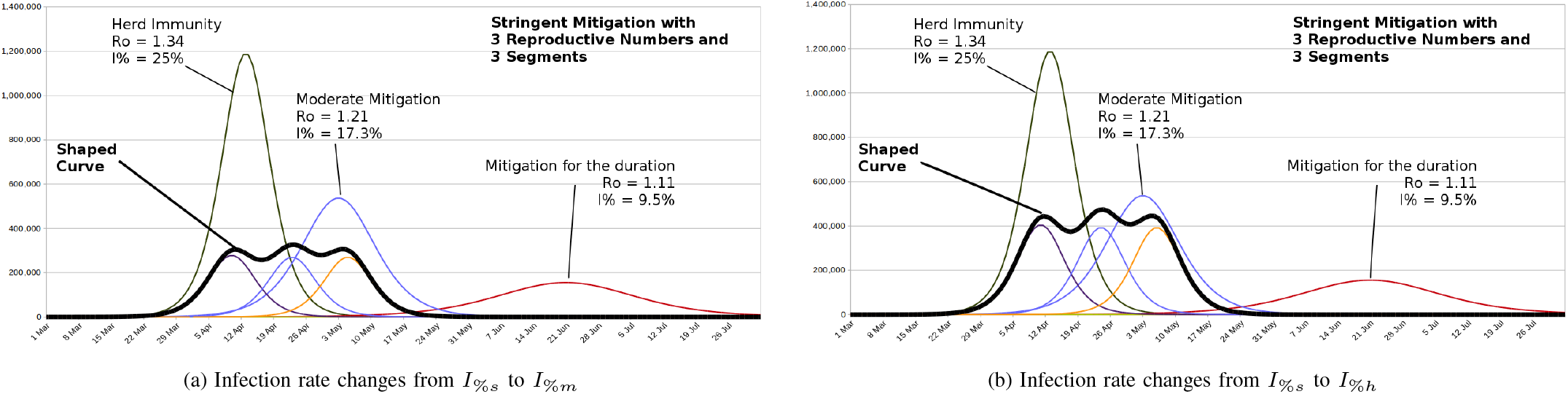
Examples for stringent mitigation measures and 3 segments showing curves for unmitigated herd-immunity, moderately mitigated, stringently mitigated, and controlled infections.

Figure 9 shows a few different implementations. In Figures 9(a) and 9(b) the timings for relaxing segment mitigation were chosen to make better use of the delay introduced by stringent mitigation. Dividing the population into 6 segments has now made it possible to reduce the amplitude of the controlled curve significantly, even when the infection rate reaches the high value of 25% in Figure 9(b). Since the controlled curve ends well before the curve with the stringent mitigation, the amplitude can be reduced further by introducing more segments.

**Fig. 9.**
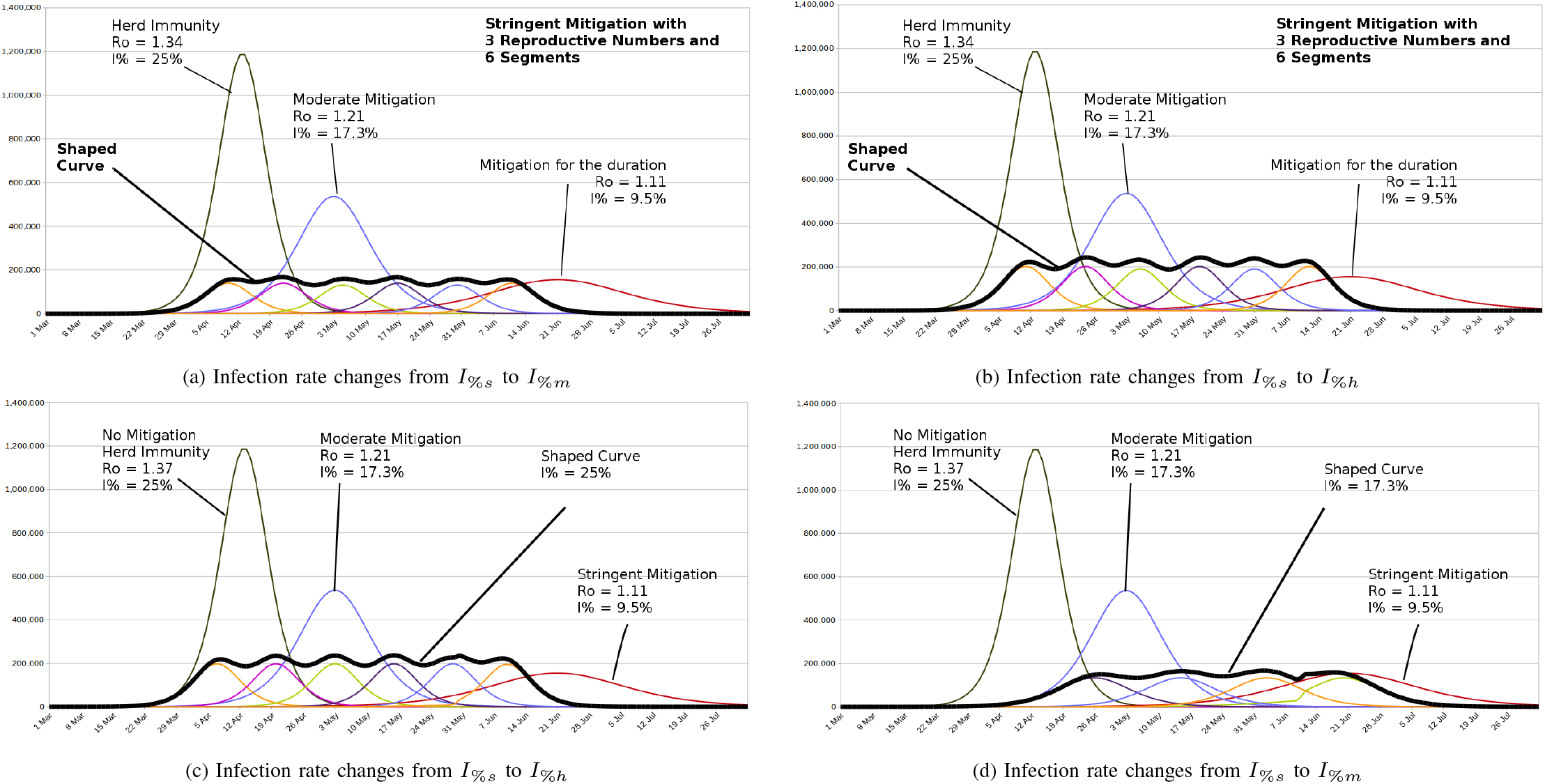
Examples for various mitigation measures and up to 6 segments showing curves for unmitigated herd-immunity, moderately mitigated, stringently mitigated, and controlled infections

In Figures 9(c) the outbreak is started without delay and this is the soonest it can be started under the same criteria as Figures 9(b), but with 4 controlled segments.

In Figure 9(d) the reproductive number is limited to *R*_*m*_ upon relaxing the mitigation for segments. This is now even more realistic for practical implementation, and notice that the start of the outbreak is automatically delayed compared to the unmitigated situation, since it cannot begin sooner than the moderate case allows. As before, the extent of the outbreak is also confined by *R*_*s*_, beyond which no control is possible.

## V. Custom Curve Programming for Changing Needs

The active pandemic curve management method is so versatile that the curve can be programmed to custom fit it to specific criteria, such as changing needs. In Figure 10(a) the curve is adapted to meet declining resources, for example to account for a decline in the health care work force due to more and more doctors and nurses falling ill to the virus. In Figure 10(b) the curve is programmed for an increase, such as a field hospital that becomes operational in the middle of the outbreak.

**Fig. 10.**
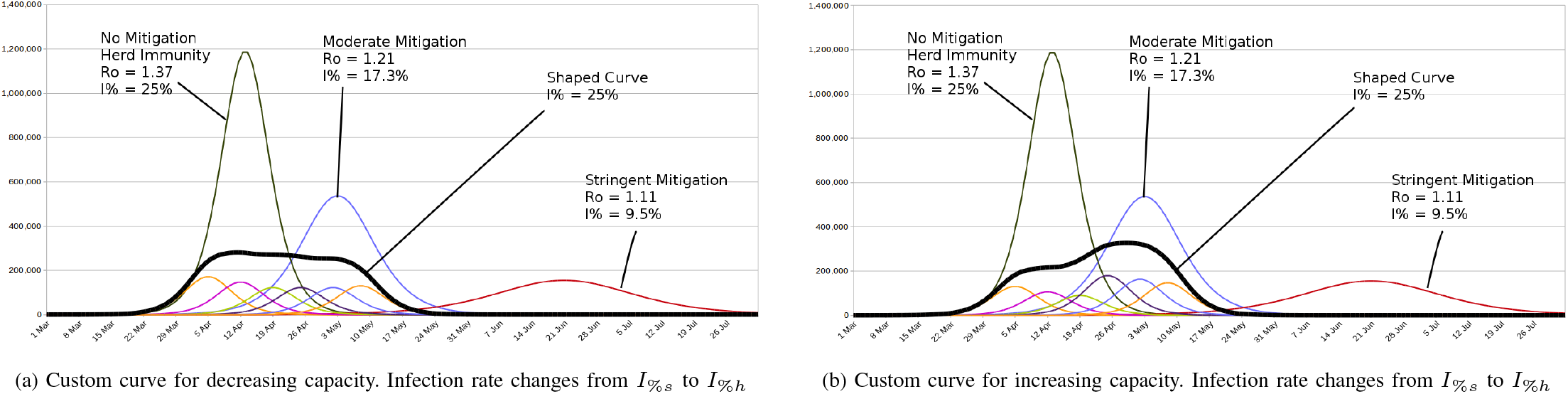
Examples of programming the curve to meet changing needs.

## VI. Discussion

The examples show great versatility in implementing MAPCM to program the curve given available mitigation. The numbers used are unimportant, and the models can be replaced by any appropriate substitute; but these examples show that moderate and stringent mitigation can be applied in a smart way to program the curve to desired needs. Mixed modes are also possible within the same strategy. In other words, it’s not necessary to limit the reproductive numbers in any way. They can be applied in any configuration to segments. Segmentation is not covered in detail in this paper for the sake of brevity, but can be done in many different ways.

### A. Origin of MAPCM

To the author’s best knowledge this methodology has never been proposed, let alone been attempted, for managing the outbreak of an epidemic. It uses a quasi-open-loop control method borrowed from electrical engineering to shape the outbreak curves. Similar pulse-shaping is performed for electromagnetic launcher technology shown in Figure 11 in the author’s laboratory during every firing event, except that it all happens within one-thousandth of a second instead of over months. There are some similarities between controlling a pandemic curve and controlling the railgun. Both are complex systems and each produces an unwanted curve that needs to be shaped into something else. Applying the method to a pandemic makes it possible to get ahead of the curve by controlling the outbreak, instead of staying behind the curve and letting a viral outbreak control a country.

**Fig. 11.**
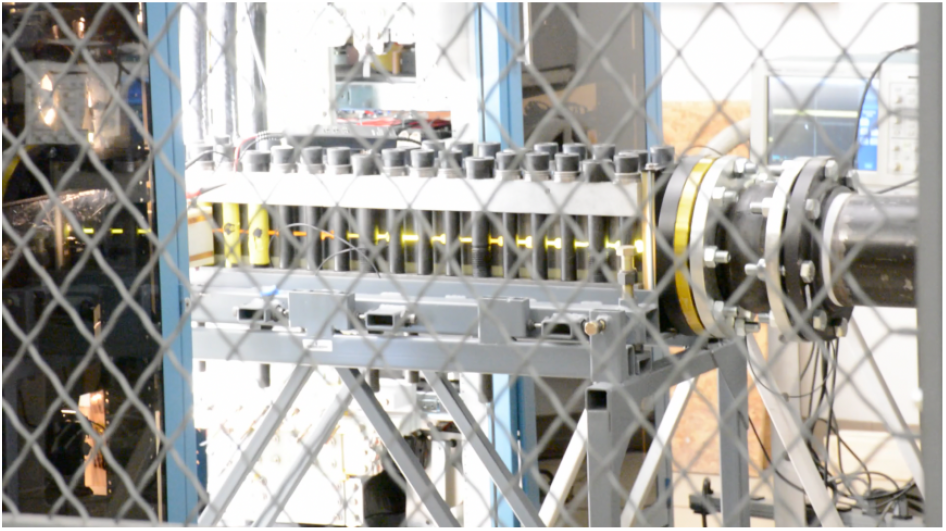
Electromagnetic launcher

The method can be useful to many countries that want to get ahead of the curve, especially those with economies that are too fragile to go into full-on lock-down like the USA has done. In the USA, the method could be implemented at the national level or at the state level. Since every locality has a unique character with its own unique challenges, the viability of this reshaping method needs to be adapted to their own models and their own mitigation measures to address their specific needs. The MAPCM method can be implemented early, before the virus has infected significant portions of a population, or later, to ease out of containment.

This method was shared with the White House Coronavirus Task Force in the beginning of March 2020 via the office of a congressman. It was also shared with a few governors by fax. However, no response was received. The author is seeking funding and potential partners to expand capabilities in this area.

### B. Opposition to MAPCM

Since this method is not intuitive, it may seem like time travel to people not familiar with control methods when applying it to disease. This is not the case. It only seems so because it involves a controlled system.

Another possible objection is that people are being infected deliberately. This isn’t the case either. Infections that would have all happened in a short period with healthcare itself incapacitated are spread out over a long period with healthcare intact.

But what if the death rate is very high? The death rate should not make any difference, because the MAPCM doesn’t sacrifice lives, but saves lives. Figure 12 plots the fatality rate among all resolved cases for the USA up until 20 March 2020, when it is 37%. However, the figure is bloated since it doesn’t include unconfirmed infections that make up a high percentage of the population. Furthermore, it should be noted that using the number of recoveries and number of deaths on a specific day in the equation:

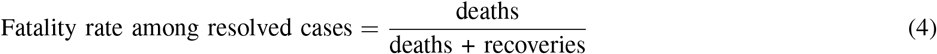

is not comparing apples to apples, because the average occupancy in days differ between patients who expire and those who are discharged from hospitals. Hospital policies before discharging a patient also differ from country to country and hospital to hospital. In Figure 12 the fatality rates among resolved cases include plots for inpatient stays that are offset by +5 and −5 days, showing that only a few days can make a big difference in calculating the death rate among resolved cases. According to [3] the difference is only 2 days.

**Fig. 12.**
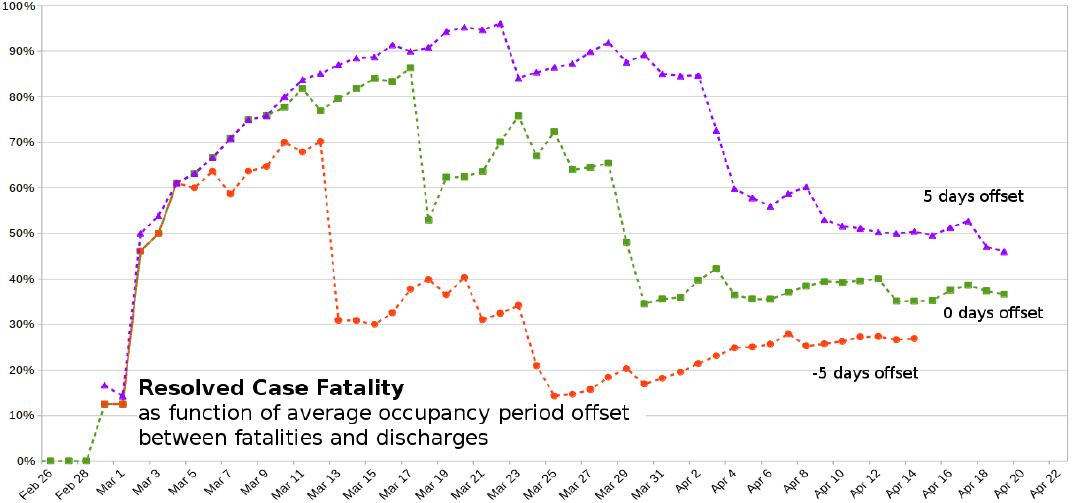
COVID-19 fatality rate among resolved cases in the USA for −5, 0, +5 day offsets between average inpatient stays of deaths and recoveries.

Using full mitigation, the lives of only some people (who are being considered essential, such as doctors and nurses, food distribution, delivery, etc.) are being risked in the war against a pandemic, while the rest of the population are forced to isolate. During a war the security of the nation is everyone’s responsibility. Rather than isolating people by force if herd immunity is not desired, the rest of the population can be educated to protect themselves the way doctors and nurses do when dealing with infected patients, and allowed back in the activated economy in a controlled manner to do their part. The level of self-isolation can then be left up to each individual to choose for themselves. Masks can be worn in grocery stores and public places. Those who are known to be at risk, such as the aged and those people with underlying conditions should obviously be protected by keeping them quarantined as much as possible.

The strategy of full mitigation in the USA is a great starting point for addressing a pandemic. However, prolonging it will put the entire nation through a lot of misery with many businesses closing and tens of millions of people losing their jobs. It is totally pointless to place a nation’s economy in a comatose state for several months when it can be surgically resolved in a controlled manner:

- MAPCM saves lives.
- The curve and the outbreak timing become programmable.
- The method allows spreading out the cases without halting the economy or sacrificing lives.
- The duration of the outbreak and mitigation can be reduced significantly compared to full mitigation.
- Herd immunity can be attained safely in a controlled fashion.
- The curve can be adjusted to accommodate changing healthcare resources, for example a field hospital becoming operational.
- The success of the method can be improved by implementing more accurate models, because control decisions are made based on predictions, however The method can be implemented even without a good model.
- The accuracy of all appropriate models can be improved by eliminating unknown variables via a scientific process of testing that has been lacking thus far. Small, representative population studies are needed for extrapolation.
- The method can be improved by a feedback loop using test data. Feedback control can handle large time delays if done correctly. The better test data is in accuracy, scale, and timeliness, the more responsive this method can be.
- The case data for the examples are based on US national averages. It would be better to implement the method at the state level, than at the national level.
- Partitioning is not limited to geography, but can be implemented based on demographics, and any number of methods.
- If those that are at risk can be isolated, such as the elderly and those with underlying issues, while the rest of the population is gradually loosened back to normal life, then the death rate can be reduced further.
- Small populations are infected faster than large populations given that everything else remains the same.

## VII. Conclusions

Sustained full mitigation for flattening the pandemic curve has severe drawbacks and causes incalculable damage to a nation’s economy. A far better way to deal with an epidemic is to take control of the outbreak and actively reshape that pandemic curve using a new technique described in this paper. For the reshaping method to be successful, there are a few objectives that need to be accomplished:

- The duration of the outbreak must be shortened and stopped in its tracks,
- the hospitalizations and critical care need to remain under the available healthcare capacity threshold,
- while allowing breathing room for the economy,
- all without putting more lives at risk.

A new technique for managing a pandemic, coined *MAPCM*, has been introduced by means of examples. It uses a quasi-open-loop control method to shape the outbreak curve for the spread of an infectious disease into a population. MAPCM makes it possible to impose controlled mitigation amid a pandemic crisis to keep the hospitalizations within available healthcare capacity thresholds while keeping the economy moving without sacrificing lives. But those are not its only features. It was shown how the amplitude and duration of a controlled curve can be traded off against one another and that the containment can be shortened in duration compared to full mitigation. It was demonstrated that the method can also be used to program the outbreak curve to meet changing needs during the outbreak. Finally, while full mitigation leaves a susceptible population open to secondary outbreaks, this method makes it possible to reach herd immunity safely and in a controlled manner.

Better results can be achieved using the method than imposing sustained mitigation by itself. MAPCM is promising for implementation early in an outbreak cycle, but also for easing out of containment. The rules for segmentation and the specific mechanical logistics for controlling the curve of an actual country or locality depend on the unique identity and character of that locality.

## Data Availability

All data is publicly available and has been widely distributed.

http://coronavirus.gov

## Appendix

### A. Definitions

#### 1) Time

So as not to be confused by the Metric System, time is not measured in seconds, but in days, for the simple reason that case data are typically collected and recorded on a daily basis.

#### 2) Reproductive Number

In this paper the *reproductive number, R*_*o*_ refers to the average number of people being infected per person in a day at the start of the outbreak as defined in equation (1). This definition has the advantage that it can be deduced directly from the data collected.

#### 3) Infection Rate

The *infection rate, I*_%_ is the percentage of a population that will be infected by the end of the outbreak cycle.

#### 4) Hospitalization Rate

The percentage of infected people who become hospitalized.

#### 5) Fatality Rate

The percentage of infected people who expire. Also called the *mortality rate* or *death rate*.

### B. The Logistic Model used in Examples

The simplest closed form analytical equation for estimating a viral outbreak with no external intervention, is according to the logistic function that gives cumulative infections:

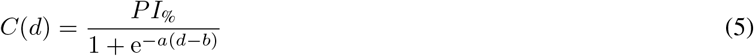

where *d* is the number of days in, *P* is the population size, *I*_%_ the infection rate, *b* a time lag, and *R*_*o*_ =≅ e^*a*^. The derivative of (5) gives the daily infections.

As noted before, this paper presents a method and not a model. The model and the numbers used in this paper were for illustration and are unimportant. The MAPCM is compatible with any other appropriate model.

### C. Estimation of Asymptomatic Infections

**Fig. 13.**
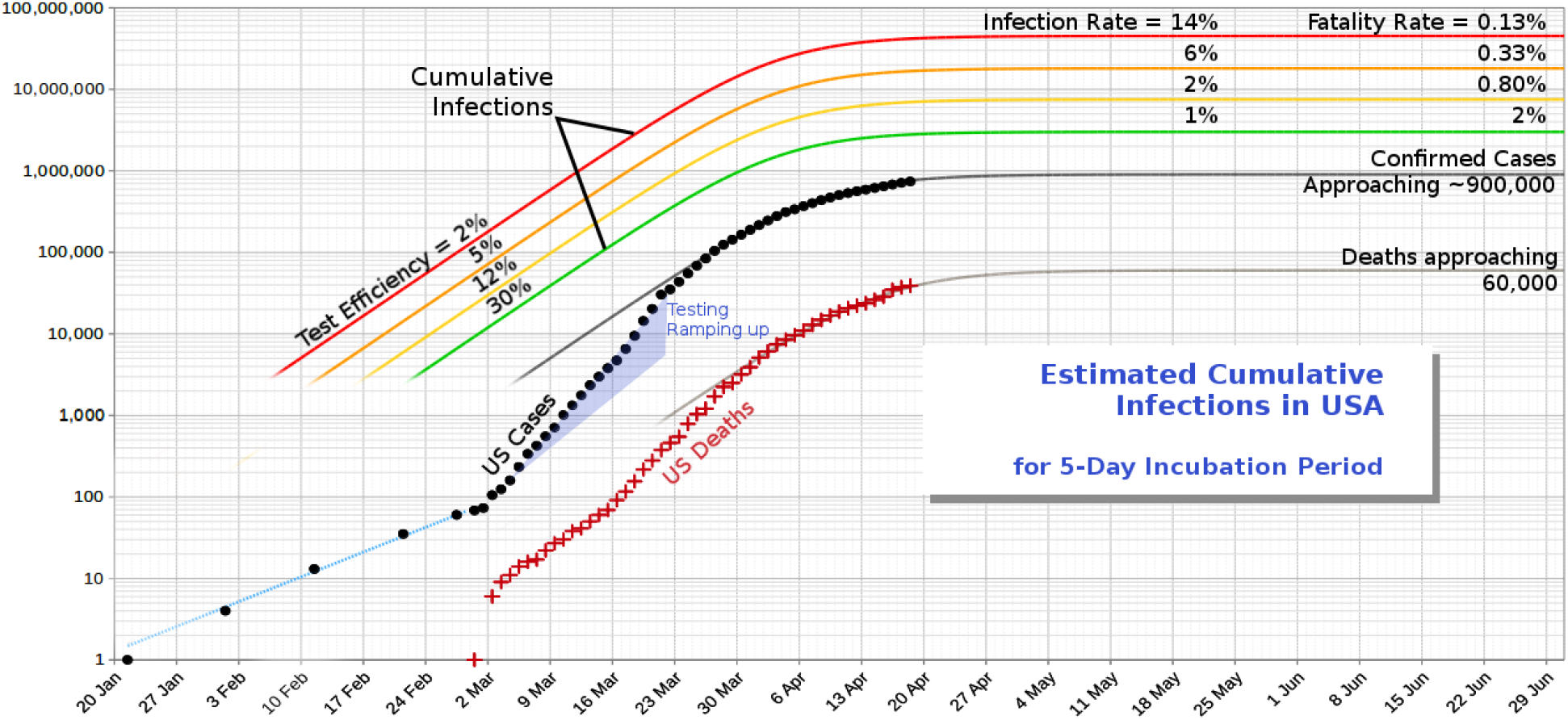
An estimate of the number of infected people in the USA and a projection of COVID-19 cases as of 19 April 2020.

The plots^4^ in Figure 13 estimate the cumulative number of COVID-19 infections in USA as function of test efficiency. An average incubation period of 5 days is assumed. *Test efficiency* is defined as the percentage of infected people who’ve tested positive for COVID-19 on average. Since the actual test efficiency, infection rate, and fatality rate will remain unknown at least until there have been sufficient population studies, lines were plotted for test efficiencies of 2%, 5%, 12%, and 30%. These plots are for the entire country.

## Footnotes

1 The number of people infected per person in a day during the initial stage of the outbreak. See Appendix for definitions.

2 Infection rate is herein defined as percentage population infected. See definitions in the Appendix.

3 The spikes in the curves are due to a derivative being taken of a logistic function with a discontinuity when *R*_*o*_ changes. Spikes like these do occur in practice also.

4 Data for confirmed cases and deaths obtained from [4]

## Notes

### Competing Interest Statement

The authors have declared no competing interest.

### Clinical Trial

There were no trials in the study.

### Funding Statement

This work was not funded

